# Prefrontal glutamate neurotransmission in PTSD: A novel approach to estimate synaptic strength in vivo in humans

**DOI:** 10.1101/2021.04.09.21255211

**Authors:** Lynnette A. Averill, Lihong Jiang, Prerana Purohit, Anastasia Coppoli, Christopher L. Averill, Jeremy Roscoe, Benjamin Kelmendi, Henk M. De Feyter, Robin A de Graaf, Ralitza Gueorguieva, Gerard Sanacora, John H. Krystal, Douglas L. Rothman, Graeme F. Mason, Chadi G. Abdallah

**Author notes:** Corresponding author*: Chadi G. Abdallah, M.D.; Menninger Department of Psychiatry, Baylor College of Medicine; 1977 Butler Blvd, E4187, Houston, TX, 77030; *Email:.

## Abstract

Trauma and chronic stress are believed to induce and exacerbate psychopathology by disrupting glutamate synaptic strength. However, *in vivo* in human methods to estimate synaptic strength are limited. In this study, we established a novel putative biomarker of glutamatergic synaptic strength, termed energy-per-cycle (EPC). Then, we used EPC to investigate the role of prefrontal neurotransmission in trauma psychopathology. Healthy control (n=18) and patients with posttraumatic stress (PTSD; n=16) completed ^13^C-acetate magnetic resonance spectroscopy scans to estimate prefrontal EPC, which is the ratio of neuronal energetic needs per glutamate neurotransmission cycle (V_TCA_/V_Cycle_). Patients with PTSD were found to have 28% reduction in prefrontal EPC (*t*=3.0; *df*=32, *p*=0.005). There was no effect of sex on EPC, but age was negatively associated with prefrontal EPC across groups (*r*=–0.46, n=34, *p*=0.006). Controlling for age did not affect the study results. The feasibility and utility of EPC were established. Patients with PTSD were found to have reduced prefrontal glutamatergic synaptic strength. These findings suggest that reduced glutamatergic synaptic strength may contribute to the pathophysiology of PTSD and could be targeted by new treatments.

**Highlights:**

- Glutamatergic synaptic strength is critical for brain function in health and disease.
- *In vivo* in human methods to estimate glutamatergic synaptic strength are limited.
- We here propose a new approach to estimate glutamatergic synaptic strength.
- The new method employs carbon-13 magnetic resonance spectroscopy (^13^C MRS).
- The utility of the new approach was demonstrated in posttraumatic stress disorder (PTSD).

## INTRODUCTION

Stressors, whether acutely overwhelming or chronic, may trigger or exacerbate posttraumatic stress disorder (PTSD) [1]. While the neurobiology of trauma and stress is not fully known [2], converging evidence suggest a critical role for glutamatergic synaptic connectivity alterations that produce the dysfunction of brain networks regulating memory and emotion associated with the PTSD symptoms [3-5]. This hypothesis is emerging from many sources of data, including preclinical and postmortem data, and indirect human *in vivo* findings from gross brain structure, functional connectivity, total neurochemical levels, or various receptors binding potentials [4, 6, 7]. Yet, a major obstacle in the field remains the lack of a more direct and dynamic assessment of glutamatergic synaptic strength *in vivo* in patients.

Here, we establish a novel metric of glutamate function that is believed to directly reflect glutamatergic synaptic strength [8]. Synaptic strength is defined as the magnitude of the postsynaptic response to a presynaptic action potential. Traditionally, changes in synaptic strength, such as long-term potentiation or long-term depression, are measured using electrophysiologic techniques [9]. However, considering the tight coupling between synaptic signaling and brain energetics [10], it is possible to infer overall glutamatergic synaptic strength within a brain region based on concurrent measurement of the rate of neuronal oxidative energy production (V_TCAn_) and glutamate neurotransmission cycling (V_Cycle_) [8]. Briefly, brain energy budget calculations indicate that cerebral signaling costs approximately 80% of total neuronal energy and that the majority of signaling energy is used on glutamate postsynaptic transmission and action potentials (∼71%) with only 9% spent on glutamate release and cycling [11, 12]. In fact, a majority of the currently available functional neuroimaging tools [e.g., functional magnetic resonance imaging (fMRI) and fluorodeoxyglucose positron emission tomography (FDG-PET)] are based on the experimental observation that brain energy metabolism is directly related to neuronal signaling [13]. One limitation of fMRI and FDG-PET is that they lack the capacity to concurrently measure the rate of synaptic glutamate release (V_Cycle_). The aim of this PTSD study is to investigate energy per cycle (EPC; i.e., V_TCAn_/V_Cycle_ ratio) as a putative biomarker directly related to glutamatergic synaptic strength [8].

Preclinical studies suggest that trauma and chronic stress reduce glutamate synaptic density, downregulate postsynaptic ionotropic glutamate receptors, and alter cortical functional connectivity reflecting overall reduction in prefrontal synaptic strength [14]. In humans, various brain imaging findings have been considered as biomarkers of this stress-related synaptic dysconnectivity [1, 15-17]. Trauma and stress-related disorders were repeatedly associated with gray matter deficits, especially in the prefrontal cortex. Reductions in cortical volume and thickness were reported in individual and meta-analysis studies [18-21]. Similarly, volumetric and shape analyses correlated trauma and stress psychopathology with gray matter deficit [22-24]. At the functional level, task and connectivity studies have identified broad circuit and large-scale brain network disturbances in trauma and stress-related disorders [4, 25, 26]. Moreover, disruption in global brain connectivity in the prefrontal gray matter was repeatedly related to the pathology and treatment of PTSD and other stress-related disorders [27-38]. At the neurochemical level, studies have investigated total levels of glutamate or binding potential of glutamate receptors and glutamate-related vesicles [6, 34, 39-44]. These approaches have numerous strengths including wide availability, good space and time resolutions, and ability to conduct whole brain assessments or study the full connectome. A main impediment to the utility of these previously identified biomarkers is the high overlap between healthy control participants and patients [45]. Another limitation is the complexity in interpreting the findings, as these measures do not specifically assess synaptic glutamate transmission but rather upstream input (e.g., glutamate receptors or vesicles) or downstream output (e.g., functional connectivity). The neuropsychiatry field may greatly benefit from establishing a biomarker that is directly related to synaptic neurotransmission, especially if this biomarker is tightly controlled in normal conditions.

^13^C Magnetic Resonance spectroscopy (^13^C MRS), combined with the stable isotope ^13^C acetate, is a specialized method to measure glutamatergic synaptic strength. ^13^C MRS allows the computation of EPC, which is the ratio of the rate of neuronal energy production divided by the rate of glutamate/glutamine cycling (V_TCAn_/V_cycle_). This ratio is equivalent to the neuronal energy consumed per glutamate cycle. EPC is a biomarker highly preserved across species and is a unique measure of glutamatergic synaptic strength [46-48]. Strength is defined as the synaptic energy consumption (in units of glucose molecules oxidized) required to support the depolarization induced by the release of one neurotransmitter glutamate molecule. These are the major energy consuming processes that contribute to the EPC ratio. Here, we used advanced methods for acquiring ^13^C MRS in the human frontal lobe [49-52], a brain region closely related to psychopathology that was not previously accessible to ^13^C MRS studies [48].

In this study, we aimed to demonstrate altered glutamatergic synaptic strength, as measured by EPC, in the prefrontal cortex of patients with PTSD, compared to healthy control. We hypothesized that the PTSD participants will present significant reduction in prefrontal EPC.

## METHODS

### Study Participants

Healthy control and individuals diagnosed with comorbid PTSD between the ages of 18 and 65 were enrolled in this study. All study procedures were approved by the Yale institution review board. All participants completed an informed consent process prior to enrollment. None of the scans from this cohort were previously reported.

Participants had no contraindication to magnetic resonance imaging, no dementia or mental retardation, no traumatic brain injury, no unstable medical condition, no pregnancy or breastfeeding, and were on a medically accepted birth control method. A negative urine toxicology test and, for women, a negative pregnancy test were required. Healthy participants were excluded if they had a lifetime history of any psychiatric disorder. Primary PTSD diagnoses were determined by a structured interview and participants were enrolled if they have: 1) been on stable serotonin reuptake inhibitor antidepressant or on no antidepressants for at least 4 weeks; 2) no diagnosis of bipolar or psychotic disorder; 3) no current substance/alcohol use disorder; 4) no current serious suicide risk; and 5) no current treatment with select medications that modulate excitatory aminoacid transporters, e.g., riluzole, ceftriaxone, pentoxifylline. Considering the high comorbidity between PTSD and depression [53] and that our research program focuses on treatments for severe PTSD cases, as expected the PTSD participants met criteria for secondary diagnosis of major depression. The PTSD checklist (PCL) and Quick Inventory of Depressive Symptomatology (QIDS) self-report of 16 items were completed to assess PTSD and depression severity, respectively [54, 55].

### ^13^C MRS Acquisition & Processing

Prefrontal ^13^C MRS acquisition and preprocessing followed our previously established procedures [8]. MRS data were acquired on a 4.0 T whole-body magnet interfaced to a Bruker AVANCE spectrometer (Bruker Instruments, Billerica, MA, USA). Subjects were placed supine in the magnet, with their head immobilized with foam. An RF probe consisting of one circular ^13^C coil (8.5 cm Ø) and two circular, quadrature driven ^1^H RF coils (12.5 cm Ø) were used for acquisition of ^13^C MR spectra from the frontal lobe (Fig. S1 A-B). Following tuning and acquisition of scout images, second-order shimming of the region of interest (ROI) was performed using phase mapping provided by Bruker.

^13^C MR spectra were acquired with a pulse-acquire sequence using an adiabatic 90° excitation pulse and optimized repetition time (offset 180 ppm, TR 6s). Nuclear Overhauser enhancement (nOe) was achieved by applying ^1^H block pulses before the ^13^C excitation pulse. ^1^H decoupling during acquisition consists of pseudo noise decoupling as described by Li *et al*. [50], to decouple the long-range ^1^H-^13^C coupling of the carboxylic carbon positions. The pseudo noise decoupling pulse has a constant amplitude and the phase of each 1.2-ms unit pulse is randomly assigned to either 0° or 180°. Following the start of [1 ^13^C]-acetate infusion, 6.5-minute blocks of MR spectra were acquired for 120 minutes (Fig. S1 C & S2).

^13^C MRS processing was conducted while blinded to the clinical data. Briefly, steady-state spectra were averaged from acquisitions after 70 minutes through the rest of the session. The steady-state spectra were analyzed with –2Hz/6Hz Lorentzian-to-Gaussian conversion and 16-fold zero-filling followed by Fourier transformation. An LC model approach was used to fit the peak areas of the labeled carbon positions of glutamate C5 and glutamine C5 (Fig. 1C), which overlapped with aspartate C4. Cramer-Rao Lower Bounds were used to estimate the quality of the individual measurements, averaging 5.8% for glutamate and 9.6% for glutamine/aspartate. The aspartate C4 kinetics closely track that of its glutamate precursor [56], thus it is considered to have the same percent enrichment as glutamate C5 and was subtracted from the combined glutamine-aspartate peak. The ^13^C-Glutamate/^13^C-Glutamine enrichment ratio was computed using peak areas of glutamate C5 and glutamine C5, (i.e., glutamate-C5/glutamine-C5 * f), where f is the ratio of glutamate/glutamine, measured by reference [57]). Then, EPC was calculated based on the relative ^13^C enrichment of Glutamate over Glutamine at steady state, as follows: V_TCAn_/V_cycle_ = [1 - (^13^C-Glutamate/^13^C-Glutamine)]/(^13^C-Glutamate/^13^C-Glutamine), where ^13^C-Glutamine and ^13^C-Glutamate represent the steady state ^13^C enrichments during the infusion of [1 ^13^C]-acetate (i.e., ∼ 70-120 min after starting infusion) [58].

**Figure 1.**
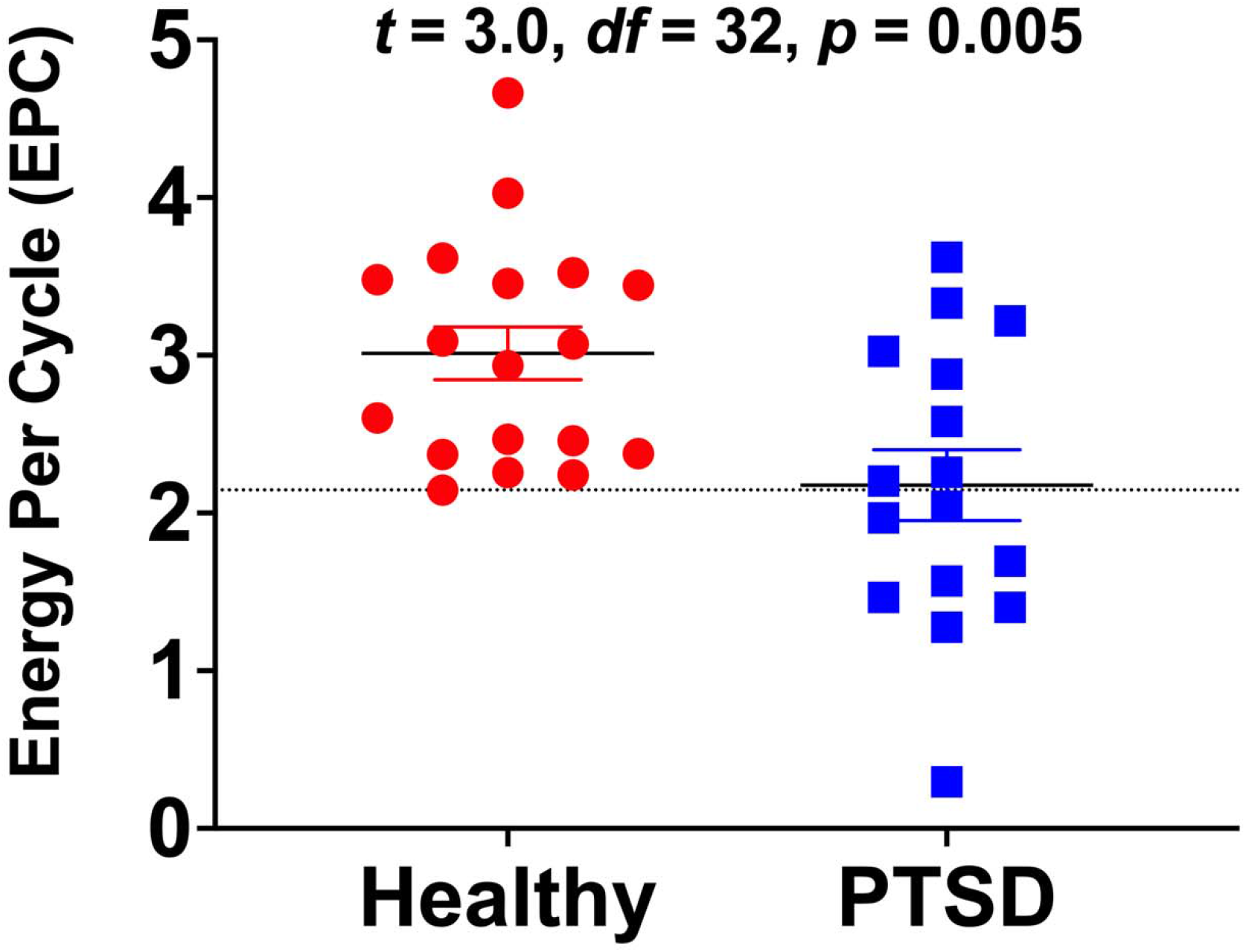
The effects of diagnosis on glutamate neurotransmission strength as measured by energy per cycle (EPC). Participants diagnosed with posttraumatic stress disorder (PTSD) were found to have 28% reduction in EPC compared to healthy controls. EPC is a measure of neuronal energetic needs (V_TCAn_) per glutamate/glutamine cycling (V_Cycle_), which is computed from the relative carbon-13 enrichment of glutamine and glutamate during steady state of [1-^13^C]-acetate intravenous infusion. The dotted line, at 2.146, marks the lowest EPC value among healthy participants. It shows that the EPC values of only 8 (50%) PTSD individuals overlapped with those of healthy control.

### Statistical Analyses

Before conducting each analysis, the distributions of outcome measure were examined. Data transformation was not needed. Estimates of variation are provided as standard error of mean (*SEM*). Considering that this is a first-in-human study, this study should be considered a first-level study implementing a novel technique, rather than a confirmatory study.

Independent t tests and chi-square tests were used to determine differences between groups. Spearman’s rank order was used for correlational analyses. Fisher r-to-z transformation was used to compare correlations between groups. General linear model examined the effects of medication status, including age as covariate. All tests were two-tailed, with the significance threshold set at 0.05. The Statistical Package for the Social Sciences (version 24; IBM) software was used for the analyses. All data will be made available through the NIMH Data Archive.

## RESULTS

A total of 34 participants (18 healthy & 16 PTSD) successfully completed the study procedures. Sex, race, age, height, and weight were not statistically different between the study groups (Table 1). Trauma exposure was not exclusionary, however, only one healthy subject was trauma exposed.

**Table 1.** Demographics and Clinical Characteristics.

|  | **PTSD (n=16)** | **Healthy (n=18)** | *p* value* |
| --- | --- | --- | --- |
| Female | 10 (62%) | 11 (61%) | 0.93 |
| White | 9 (56%) | 11 (61%) | 0.77 |
| Age (years) | 39.5±3.3 | 34.3±2.9 | 0.25 |
| Height (inches) | 65.7±1.0 | 65.3±0.8 | 0.60 |
| Weight (lbs.) | 169±12 | 148±8 | 0.16 |
| PCL | 45.4±3.9 | 1.8±0.7 | < 0.01 |
| QIDS | 14.5±0.9 | 1.7±0.4 | < 0.01 |

\* Chi-square and independent t tests were used to compare groups. *Abbreviations:* PTSD = posttraumatic stress disorder; PCL = PTSD checklist; QIDS = Quick Inventory of Depressive Symptomatology.

We first investigated the effect of group on EPC. We found a 28% reduction of EPC in PTSD (mean±SEM = 2.2±0.2) compared to healthy control (mean±SEM = 3.0±0.2; *t* = –3.0, *df* = 32, *p* = 0.005; Fig. 1). Next, we examined the effect of sex on EPC, which showed no EPC differences between males (mean±SEM = 2.6±0.2) and females (mean±SEM = 2.7±0.2; *t* = –0.3, *df* = 32, *p* = 0.75). However, there was a significant negative correlation between EPC and age (*r* = –0.46, *n* = 34, *p* = 0.006; Fig. 2). Comparing the EPC-age association between study groups showed no statistically significant difference in PTSD (*r* = –0.41) compared to healthy control (*r* = –0.49; *z* = 0.27, *p* = 0.79). Considering the relationship between age and EPC, we conducted a general linear model analysis examining the effect of diagnosis controlling for age. The general linear model showed significant group effects (F_(1,31)_ = 7.3, *p* = 0.01), indicating reduced EPC in PTSD compared to healthy control, covarying for age.

**Figure 2.**
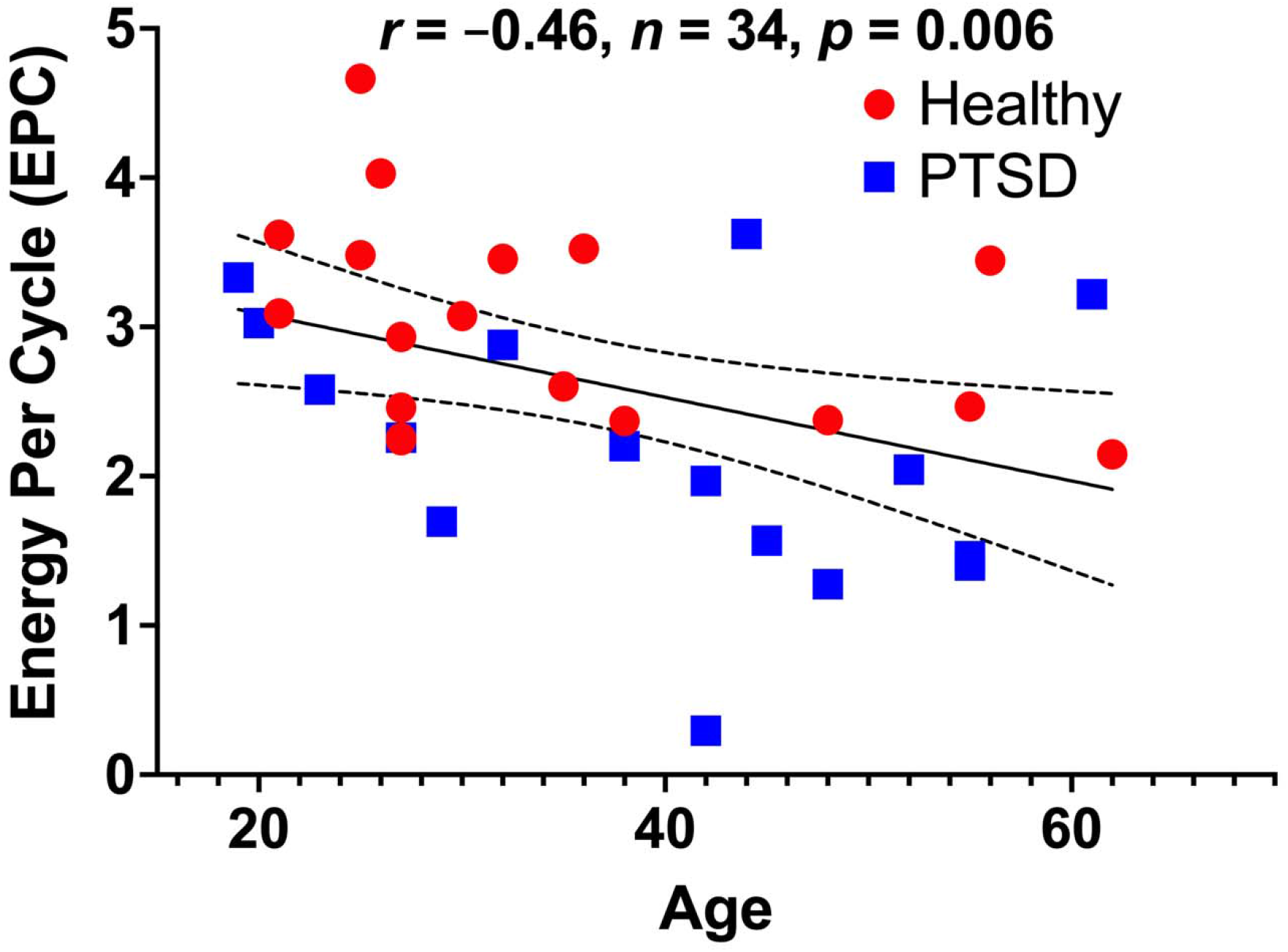
The association between age and glutamate neurotransmission strength as measured by energy per cycle (EPC). There is a significant negative correlation between age and EPC in the full cohort, as well as in the healthy and patient groups considered separately. EPC is a measure of neuronal energetic needs (V_TCAn_) per glutamate/glutamine cycling (V_Cycle_), which is computed from the relative carbon-13 enrichment of glutamine and glutamate during steady state of [1-^13^C]-acetate intravenous infusion. Abbreviations: PTSD = posttraumatic stress disorder.

## DISCUSSION

The study established the feasibility and methods for determining EPC in the prefrontal cortex *in vivo* in humans using [1 ^13^C]-acetate MRS, a relatively less complex and less burdensome approach than using ^13^C-glucose. The results also provided the first *in vivo* evidence of reduced prefrontal EPC in trauma-exposed individuals with PTSD, suggestive of a reduction in glutamatergic synaptic strength in this patient population. The data provided supportive evidence about the utility of EPC as measured by [1 ^13^C]-acetate MRS. In particular, the values of EPC in healthy participants overlapped with only 8 (i.e., 50% of) PTSD individuals, indicating that EPC is a tightly controlled biomarker in normal conditions. Together, these data underscore the role of the EPC biomarker in the neurobiology and treatment of trauma and stress-related psychopathology.

The use of EPC as measured by ^13^C-acetate MRS could hold great promise as a powerful translational treatment target biomarker. **1)** The relationship between neuronal energetics (V_TCAn_) and glutamate cycling (V_cycle_) is comparable among rodents and humans [47]. This could be highly useful during early stages of drug development, as pharmacoimaging paradigms established in rodents could be readily translated to humans. **2)** EPC is stable across differing level of neuronal activation and brain state, maintaining on average an approximately constant ratio in anesthetized, asleep, and awake brains [10, 47]. This overall stability across brain activity states is a major strength as biomarker, which simplifies acquisition paradigms and could reduce potential state dependent confounding effects across studies. **3)** The EPC ratio was previously related to psychopathology [48]. **4)** Another advantage is that in ^13^C-acetate MRS, EPC measure is calculated based on the relative ^13^C enrichment of Glutamate over Glutamine during isotopic steady state as opposed to a lengthy dynamic time course in the magnet. Together, these characteristics will ensure the rigor and reproducibility of the biomarker, as studies targeting EPC would (a) only need to acquire scans during steady state infusion of acetate, without the need for 2h acquisition to capture the full time-course of acetate kinetics. (b) The measure would not necessarily require sophisticated kinetic modeling or various input functions (e.g., plasma enrichment). (c) As a ratio of 2 metabolites acquired concurrently, it also does provide an optimal internal reference that obviates the need for common MRS correction methods (e.g., phantom replacement, tissue composition, etc.). (d) Given its dependence on steady state only, it may also permit the exploration of various routes of acetate administration instead of the intravenous infusion route. (e) Signal to noise is also optimal considering the high level of ^13^C enrichment during steady state and the fact that it is an average of long acquisition (∼ 1h in the current study). Together, these strengths of the EPC measure could significantly reduce the complexity of ^13^C MRS acquisition and facilitate its deployment at large scale if this biomarker was confirmed to be of clinical utility.

Overall, the findings of the current study highly support the use of EPC as complimentary biomarker to assess the role of glutamatergic synaptic strength in the pathophysiology of trauma and stress-related disorders. As a ratio of V_TCA_/V_Cycle_, EPC measured by ^13^C-acetate MRS does not differentiate between reduction in V_TCA_ or increase in V_Cycle_, and vice versa. However, this limitation is also a major strength of the biomarker as it is also a ratio of glutamate/glutamine ^13^C enrichment at steady state, providing ideal internal reference as well as optimal signal to noise. In our previous studies, we used ^13^C-glucose MRS to measure V_TCA_ and V_Cycle_ independently [8, 48]. Yet, we found the EPC ratio (i.e., V_TCA_/V_Cycle_) to be the most salient biomarker. In one ^13^C-glucose MRS study, we found a 26% reduction in occipital EPC in depressed compared to healthy control [48]. In a separate study, we found that the rapid acting antidepressant ketamine significantly increased prefrontal EPC in healthy and depressed participants, as indicated by its differential effects on glutamate and glutamine enrichment [8]. These latter findings were consistent with preclinical data showing differential effects of ketamine on prefrontal glutamate and glutamine enrichment [59, 60]. Finally, our previous data correlated prefrontal EPC with the psychotomimetic effects of ketamine, suggesting that EPC is not only relevant to antidepressants and stress-related psychopathology but also perhaps to psychosis mechanisms [8].

Another limitation of EPC is the lack of spatial resolution with the current ^13^C MRS methods, which are limited to large single cortical ROI and do not permit localization to a specific brain region, e.g., anterior cingulate. However, the cortical ROI targeted in this study (Fig. S1) is believed to play a critical role in PTSD psychopathology. In addition, based on postmortem and preclinical data, the glutamate abnormalities appear to be widespread throughout the prefrontal cortex [14, 61]. Our colleagues are currently developing novel ^1^H-^13^C MRS approaches that will provide higher resolution as well as access to deeper brain structures, which could be used in future studies [62]. Finally, while we demonstrated the utility of EPC in patients with PTSD, future larger studies are still required to determine the effects of antidepressants and the specificity of EPC alterations to PTSD, trauma exposure or the comorbid depression.

## Conclusion

The current report provides the logical intuition for computing glutamate synaptic strength *in vivo* in humans and details the methods for acquiring ^13^C-acetate MRS in the prefrontal cortex and for estimating EPC. It briefly describes the EPC alterations in PTSD and discusses the strengths and limitation of EPC as measured by ^13^C-acetate MRS. Overall, the results of this study support the glutamate synaptic dysconnectivity models of trauma and stress-related psychopathology [1, 6]. It shows 28% reduction in prefrontal EPC in PTSD, with a limited overlap between patients and healthy individuals (Fig. 1). These findings are comparable to previous findings in occipital EPC measured by ^13^C-glucose MRS [48], suggesting widespread cortical disruption in EPC with tightly controlled EPC values in normal condition. Another finding is the negative correlation between age and EPC, raising the possibility that the observed reduction in cortical EPC might be consistent with a phenomenon of accelerated aging in trauma and stress-related disorders [63]. Future studies should investigate the effect of antidepressants on EPC and examine whether the prefrontal EPC disruption is related to trauma exposure, to the PTSD severity and comorbidities, or to both.

## Data Availability

Data will be made publicly available through the https://nda.nih.gov/

## ACKNOWLEDGMENTS

The authors would like to thank the individuals who participated in these studies for their invaluable contribution.

## AUTHORS CONTRIBUTION

Conceptualization, C.G.A., G.S., J.H.K., D.L.R. and G.F.M.; Methodology, C.G.A., H.M.DF., R.A.dG., D.L.R. and G.F.M.; Data Curation: C.G.A., L.A.A., L.J., P.P., A.C., C.L.A., J.R., B.K., and G.F.M.; Formal Analysis, C.G.A. and R.G.; Investigation, C.G.A., L.A.A., L.J., P.P., A.C., C.L.A., J.R., B.K., R.G., G.S., J.H.K., D.L.R. and G.F.M.; Writing – Original Draft, C.G.A.; Writing – Review/Edit, all authors; Funding Acquisition, C.G.A., J.H.K. and G.F.M.; Resources, C.G.A., L.A.A. and G.F.M.; Supervision, C.G.A. and G.F.M.

