## Supplementary material for "Prefrontal glutamate neurotransmission in PTSD: A novel approach to estimate synaptic strength in vivo in humans": Fig. S1

SUPPLEMENTS

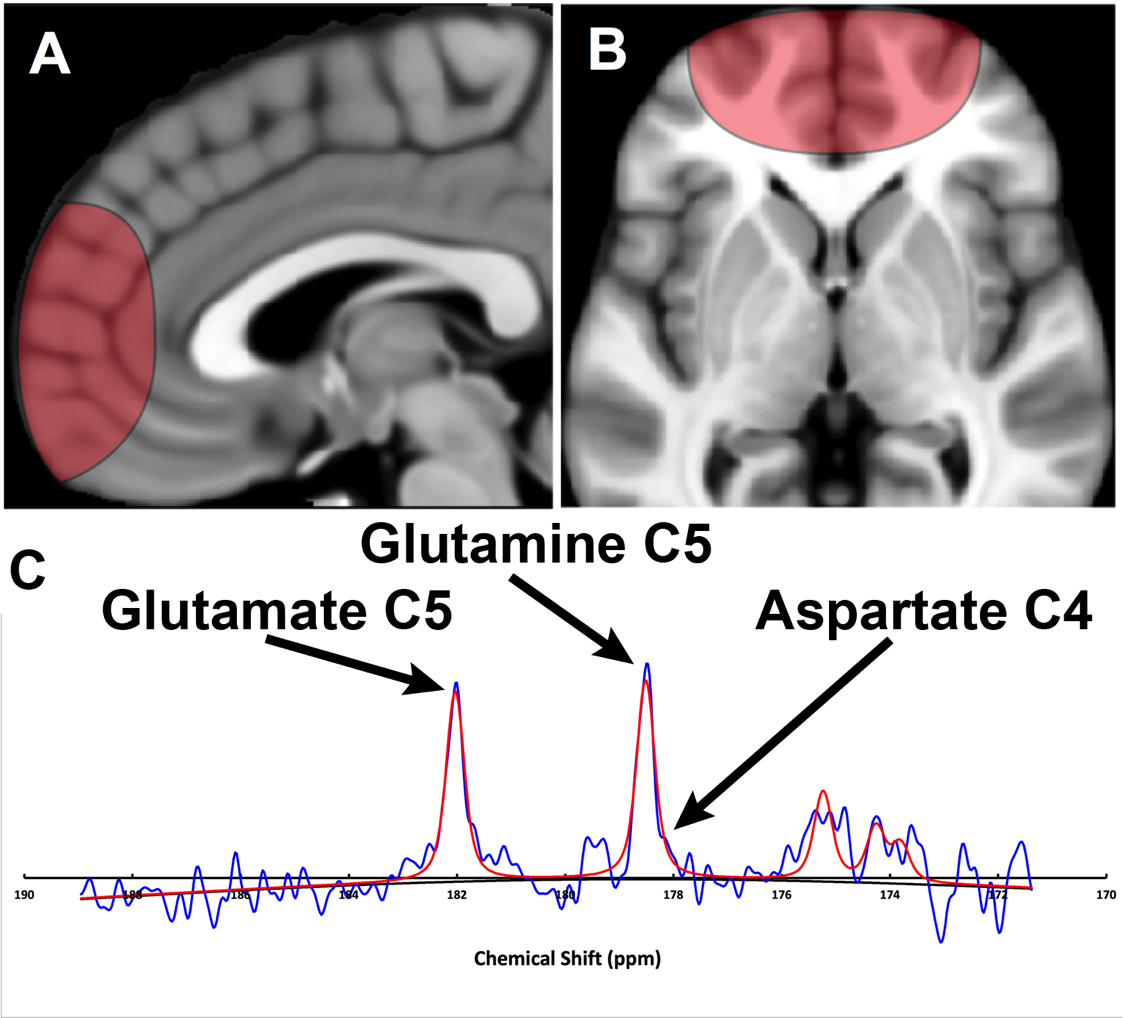

**Figure S1. Prefrontal ^13^C magnetic resonance spectroscopy (MRS) acquisition and ^13^C spectrum.** Sagittal (A) and axial (B) view of the region of interest – based on the radius of the carbon coil – primarily rostral Brodmann Area 10. (C) ^13^C magnetic resonance spectrum acquired at 4T from the prefrontal region of a study participant during infusion of [1 ^13^C]-acetate. Color code: blue = raw; red = fitted.

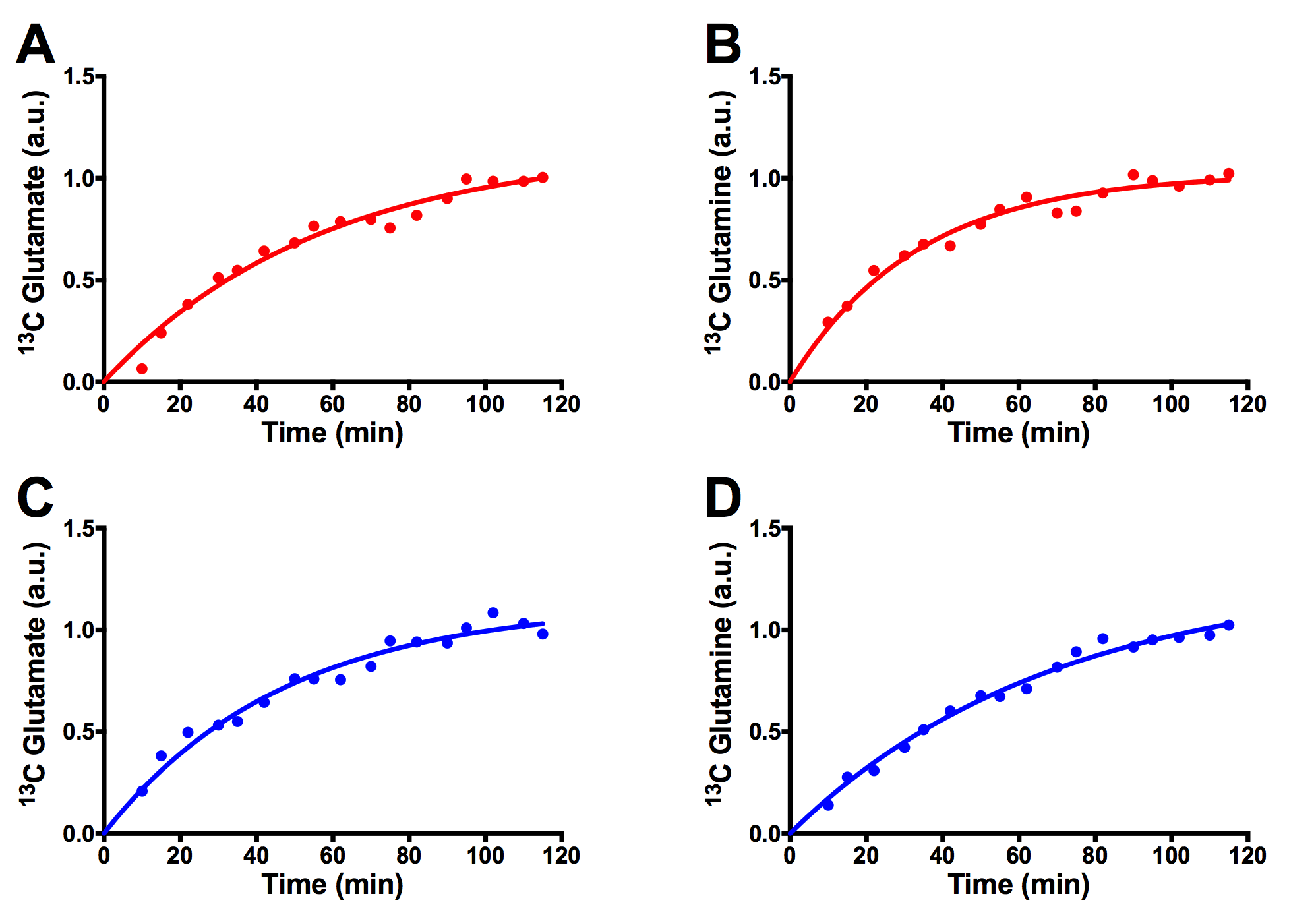

**Figure S2. Example time course of prefrontal glutamate C5 and glutamine C5 enrichment from two study participants (AB & CD).**
